# Modalities of mobile dental units worldwide: a scoping review

**DOI:** 10.1101/2025.06.01.25328767

**Authors:** Edmundo Duarte, Anderson Kaian de Lima, Sophia Marques, Angelo Roncalli

## Abstract

The strategy of transporting health teams to remote areas using equipped vehicles is effective in expanding health coverage. However, the viability of this service depends on geopolitical factors, the health needs of the population, and existing health models, and knowledge about how this modality works is essential for its successful implementation. The aim of this study is to map scientific evidence on the strategies employed by mobile dental units around the world and to establish relationships between the strategies and their effectiveness. This scoping review followed the methodological guidelines of the Joanna Briggs Institute. The PubMed, Lilacs, Scopus, Embase, and gray literature databases were searched using predefined descriptors from the Health Sciences Descriptors and Medical Subject Headings. Studies were identified and evaluated using blinded peer review to answer the guiding question: “what are the modalities of mobile dental units around the world?”. Studies without language and time restrictions, from public and private domains, that described mobile oral health care systems were included. After screening 2,059 studies and removing duplicates, 55 were included. The studies were published in 19 countries between 1974 and 2024, with observational studies and experience reports predominating. Public sources and non-profit institutions funded the majority of studies, and vans and trailers were the most common vehicles, with services provided primarily to children and vulnerable populations. Common services included dental restorations, extractions, and oral health education. Data analysis indicated that mobile dental units are a crucial component in expanding access to dental care.

## INTRODUCTION

The World Health Organization reaffirmed in its World Health Report (2008) that developing strategic health systems grounded in primary healthcare is the main approach to achieving sustainable improvements in population health (1). Patient access to health services depends on multiple factors, helping or limiting this access, and the presence of a structured health system allows the development of different levels of attention (2). The primary health attention is marked by the access to territories and health needs of the population (3). For instance, the use of specialized and emergency dental services is determined by social, epidemiological, and health service organization factors with different patient-centered structures, establishing access inequalities based on the scope of services (4).

In this context, health strategies are developed to overcome access barriers and ensure the continuity of dental care (5). Mobile health services are an effective tool for promoting oral health education, patient autonomy, and critical knowledge about risk factors for oral diseases (6). California, in collaboration with the University of Southern California, offered mobile dental units (MDU) to provide risk-prioritized dental evaluation and care for underprivileged children in 70 communities since 72% of the children did not attend a dentist in the last 12 months (7). They offered prevention, topical fluoride, and oral health education, and each child received approximately 2.9 sealants and 4.2 restorations at the of the treatment.

MDU have expanded beyond remote areas, providing care for the homeless (8) and participating in programs for pregnant women to ensure continuity of pregnancy without intercurrences (9). Considering that individualized approaches according to health needs and local characteristics are important for the efficiency of this strategy, various modalities of MDU have been developed. In riverside areas, for example, mobile river units often face environmental challenges, such as river flooding, drought, or boat inaccessibility, requiring the improvement of the strategy (10). Also, experiences are only partially positive; in Africa, MDU are associated with low levels of adherence due to patient absence (11).

Integration of MDU with efficient access to services allows the provision of oral healthcare (12). Also, literature has highlighted the planning and evaluation phases for successful implementation of these public policies and the influence of context and decision-making in outcomes (13). Evaluation metrics can encompass a range of factors, including the establishment of goals (14), monitoring of the developed activities by evaluating access (15), percentage of educational activities (6), and proportion of services provided in the integrated territories (16).

Although studies have reported the use of MDU (17), further studies are needed to examine the efficacy of this strategy on a global scale. Thus, this review aimed to map evidence on modalities of MDU worldwide and develop a theoretical model to identify the characteristics that motivate the inclusion of this service in healthcare. Also, it aimed to perform a systematic search on strategies used by the MDU modalities, establishing a relationship between strategies and characterizing their effectiveness, population coverage, period, and territory of operation.

## METHODS

### Research question

A scoping review was conducted between April and August 2024. This design was appropriate for the study, considering the limited nature of the literature. The main question of this scoping review was “what are the modalities of MDU worldwide?”, guided by the following sub-questions: “is there a specific strategy for implementing these services?”; “what is the patient profile covered by the strategy?”; “what is the geopolitical profile of the covered areas?”; “what is the timeframe for establishing the strategy?”; “which modality is implemented? Is there a justification?”; “are any characteristics of the country relevant to the use of the strategy?”; “is there an evaluation of efficiency or satisfaction?”; “what instrument is used?”; and “how many units are present in the territory?”.

### Evidence search

The research protocol for this review can be accessed on the Open Science Framework portal. The PubMed, Lilacs, Scopus, Embase, and grey literature (i.e., Google Scholar, Brazilian Digital Library of Theses and Dissertations, Catalog of Theses and Dissertations of the Coordination for the Improvement of Higher Education Personnel, and Open Access Theses and Dissertations) databases were used. A search strategy was conducted using predefined terms from Health Sciences Descriptors (DeCS) and Medical Subject Headings (MeSH) to identify studies characterizing the use of MDU in different countries. Published studies were considered, without temporal or language restrictions, as these aspects do not compromise the research question. Chart 1 presents the search strategy.

**Chart 1.**
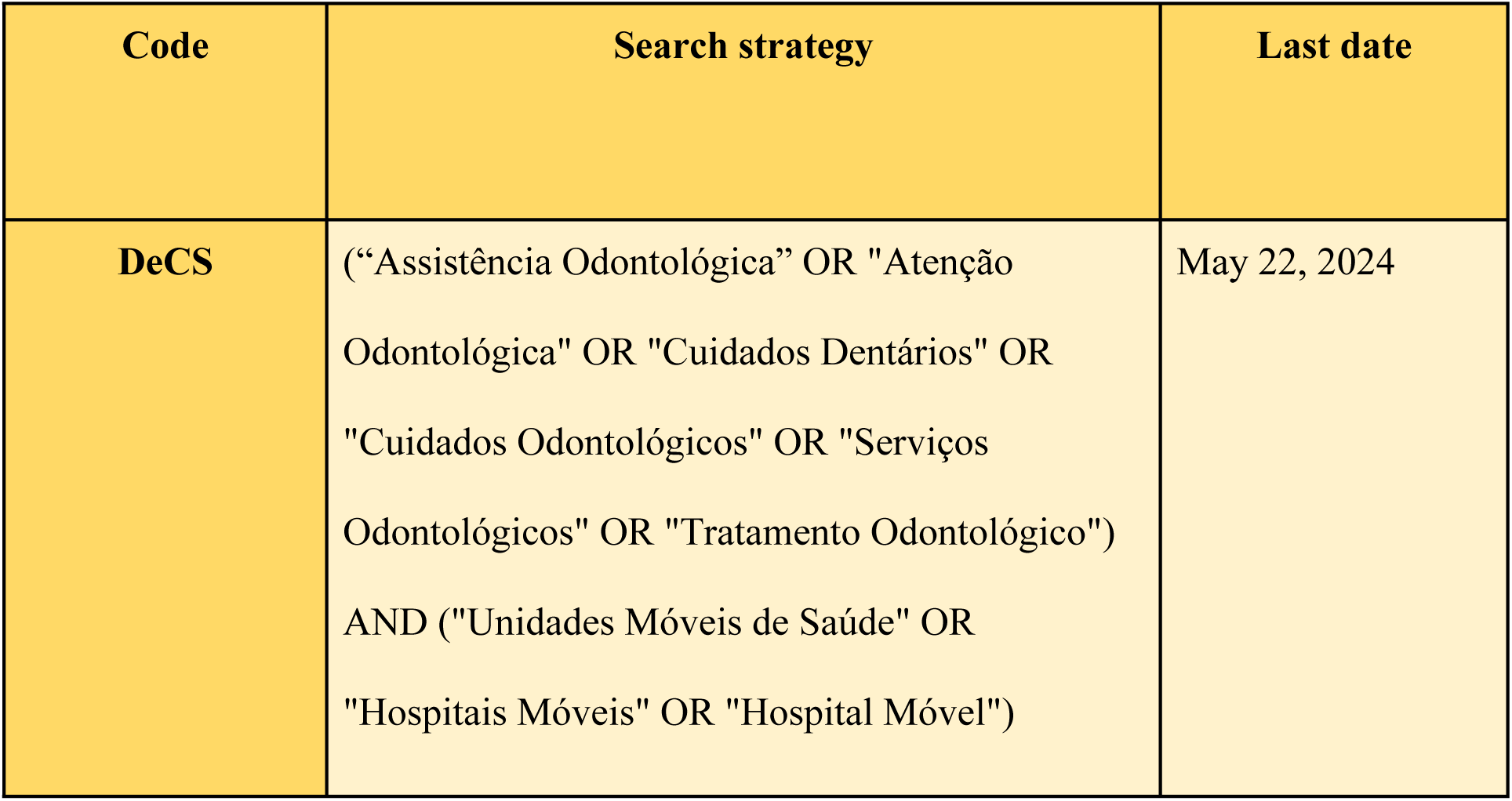
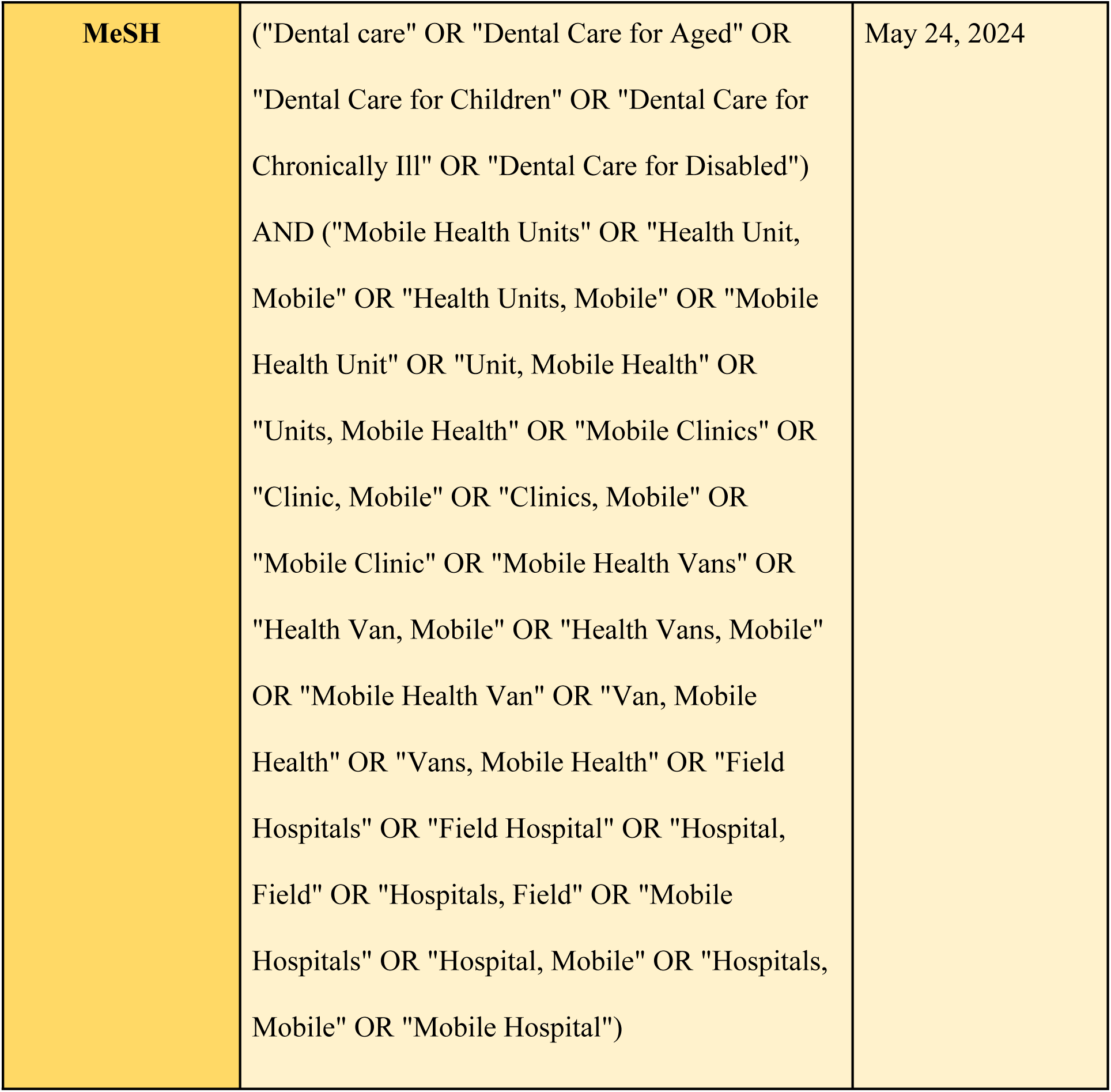
Descriptors and search strategies employed for studies retrieval.

### Study selection

The studies identified in databases were exported to Rayyan® (i.e., a reference management software developed by the Qatar Computing Research Institute) for duplicate removal, paired and blinded selection, and screening.

Titles and abstracts were initially reviewed to identify studies relevant to the goals of this scoping review and inclusion and exclusion criteria. In this phase, two independent reviewers (EDM and AKLM) evaluated the relevance of each study based on the title and abstract according to the predefined inclusion and exclusion criteria. The following studies were included: without language and time restriction, from public and private domains, and using various designs (e.g., primary and secondary studies, experience reports, and government documents). Studies addressing mobile services outside the scope of dentistry, individual reports, or incomplete texts were excluded.

In the second phase, full-text reviews were conducted for the selected studies according to the eligibility criteria, and data were extracted for analysis. A third reviewer (SQMS) resolved any disagreement between reviewers.

Two independent reviewers (EDM and AKLM) extracted data using Google Forms, a previously tested platform for this action. Two tables were created for each identified material and subsequently integrated into one table by consensus between the two reviewers. Data were mapped using the Joanna Briggs Institute guidelines. The extraction table included the following information: title, author, publication type, country of origin, institution, year of publication, aim, study design, sample size (when applicable), and main results related to research questions.

### Summarizing results

The extracted data were categorized and analyzed to answer the following research questions: 1) What are the modalities of MDU worldwide?; 2) Is there a specific strategy for implementing these services?; 3) What is the patient profile covered by the strategy?; 4) What is the geopolitical profile of the covered areas?; 5) What is the timeframe for establishing the strategy?; 6) Which modality is implemented? Is there a justification?; 7) Are any characteristics of the country relevant to the use of the strategy?; 8) Is there an evaluation of efficiency or satisfaction? What instrument is used?; and 9) How many units are present in the territory?. A narrative and descriptive structure with diagrams and tables was developed based on study analysis to align with the characteristics of the included studies and address the research questions.

## RESULTS

### Study selection

The initial search resulted in 2,059 studies, which were reduced to 1,542 after removing duplicates. Screening of titles and abstracts resulted in 100 studies selected for full-text review, with 55 being relevant to the scope of the study (Figure 1). The results were compiled to answer the predefined sub-questions and explain the MDU scenario.

**Fig 1.**
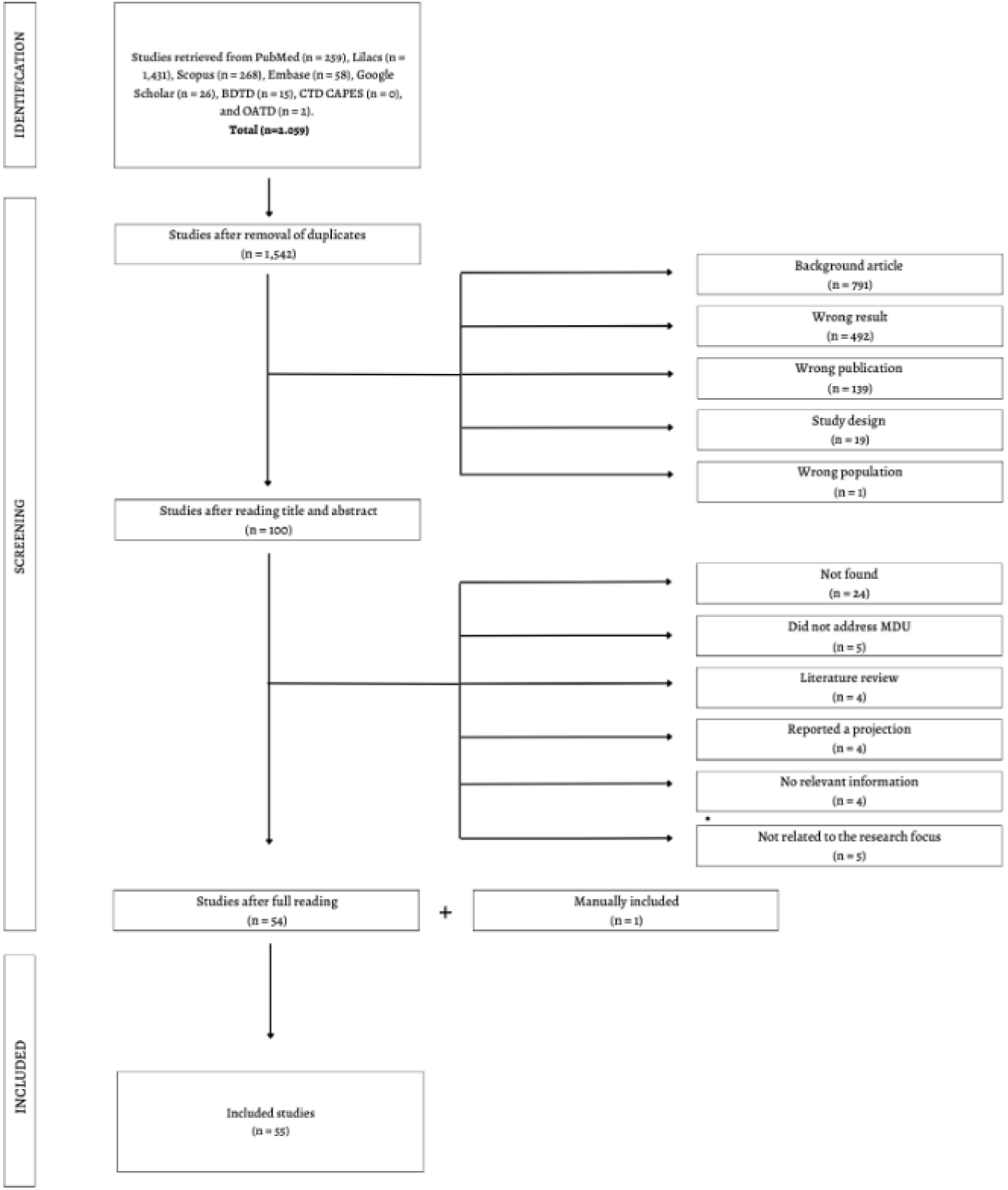
Flowchart of studies selection to characterize MDU worldwide. MDU: Mobile Dental Units; BDTD: Brazilian Digital Library of Theses and Dissertations; CTD CAPES: Catalog of Theses and Dissertations of the Coordination for the Improvement of Higher Education Personne; OATD: Open Access Theses and Dissertations. Source: the author, 2024.

When data was missing or incomplete, efforts were made to contact the authors to obtain important unpublished information. The authors were contacted via email up to three times, with one-week intervals between each attempt, whenever additional information was required.

After analyzing the results, the data were visually reorganized by creating a figure on the Canva® platform and in the Adobe Illustrator® program.

### Study characterization

All selected studies were written in English and originated from the following countries: Afghanistan (1), Australia (1), Brazil (1), Belgium (1), Slovakia (1), Spain (1), France (1), Israel (1), Malaysia (1), Peru (1), Sweden (1), Austria (1), Canada (2), India (3), Thailand (3), South Africa (3), England (6), and United States (26) (Figure 2). Publication years ranged from 1974 to 2024, with 37.5% of studies published in the last 10 years. Regarding study design, 27 were observational, 17 were reports of experience, 6 were retrospective cohort, 4 were prospective cohort, and 1 was an intervention.

**Fig 2.**
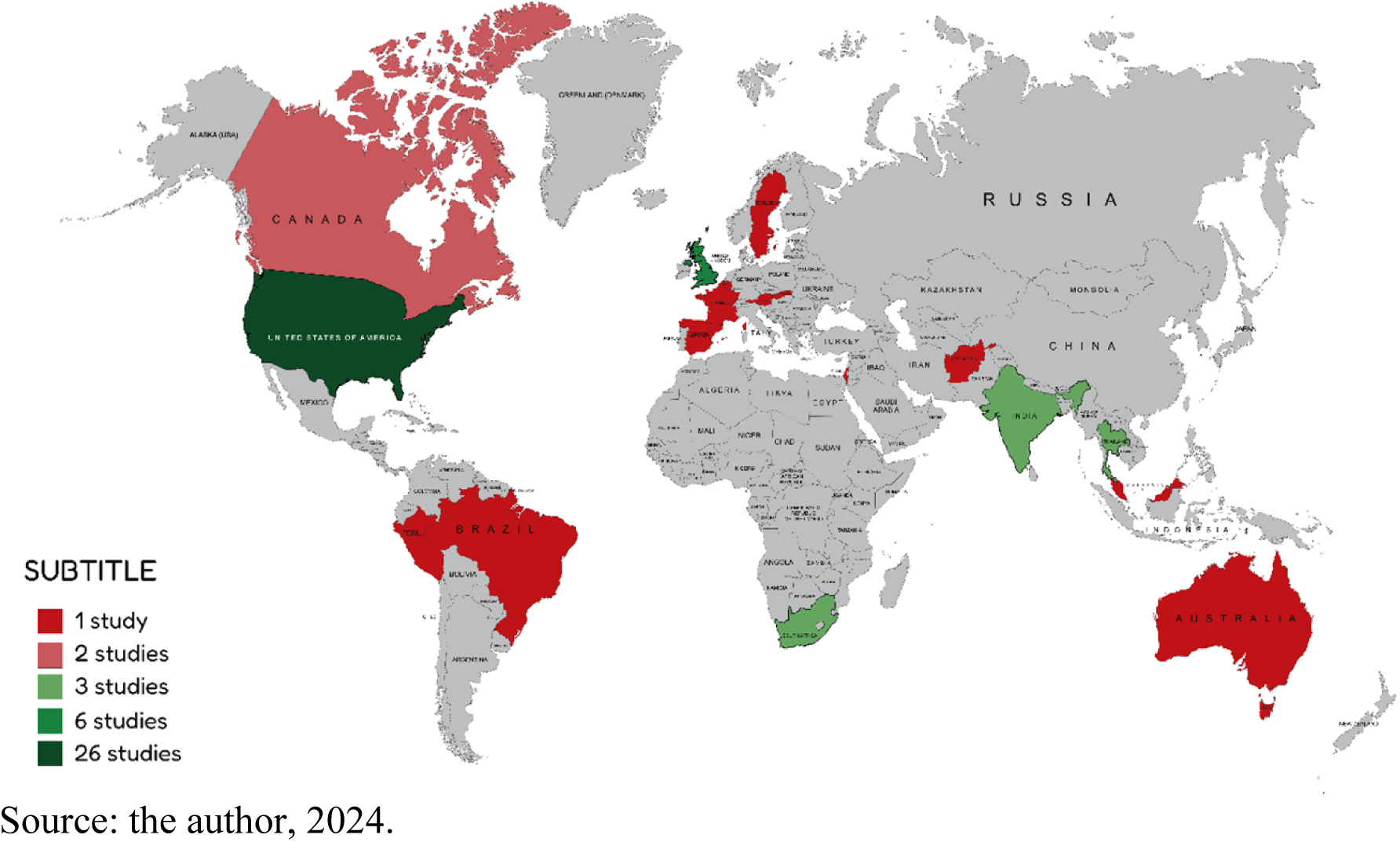
Illustration of the worldwide distribution of studies on mobile dental units.

**Fig 2.**
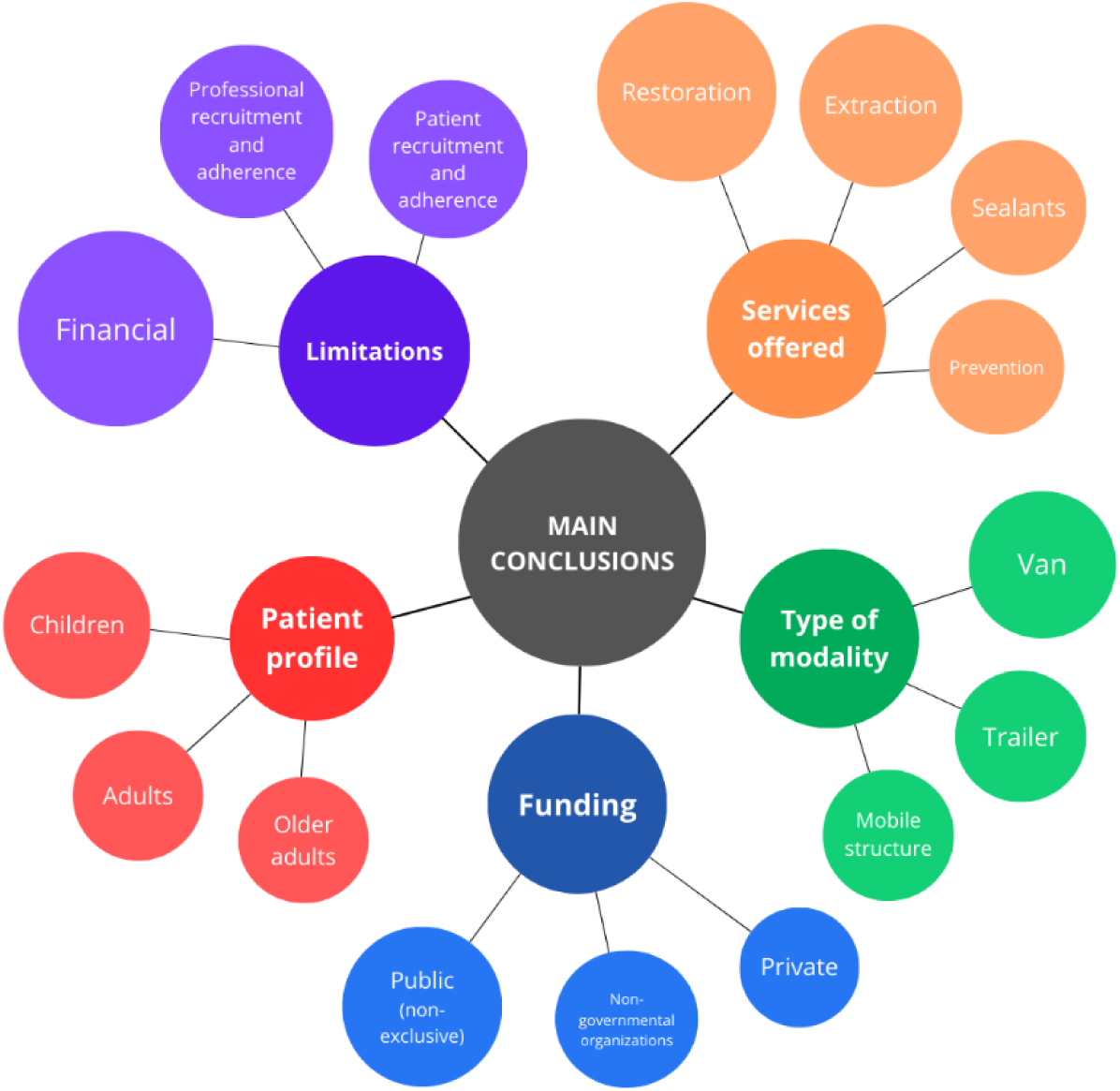
Flowchart with main findings. Illustration of summarizing results. Source: the author, 2024.

Public funding at the federal, state, municipal, or community level was the most cited (82%). Non-profit institutions, mostly supported by philanthropy, accounted for 64% of resources, followed by private initiatives (32%), patient self-funding (25%), and university programs (17%).

Most studies did not specify the modalities of transport used for MDU (41%), while others reported vans (21%), trailers (16%), and buses (9%). Other modalities included trains (5%), trucks (5%), aircraft (1%), and river vessels (1%). Approximately 9% of programs did not use vehicles to store equipment but operated with a mobile clinical structure across remote areas.

Regarding patient profiles, 31 studies (55%) focused on children in primary school, living in remote areas, or without a health system. Also, services were aimed at the following populations: socioeconomically disadvantaged, marginalized (e.g., homeless, LGBTQIA+, abuse survivors, and substance users), with disabilities or comorbidities, older adults, military, and Indigenous.

Considering the 38 studies that provided detailed information regarding services offered by the MDU, most of them (73%) reported restoration of decayed teeth using amalgam or composite, followed by extractions (65%), sealants (47%), oral health education (39%), and scaling and root planing (39%).

Although most studies originated from the United States, this review reported experiences from diverse countries across all continents, each with unique applications and particularities. Figure 2 and Frame 1 presents the main findings for each sub-question.

**Frame 1.** Summary of main findings in subcategories.

| Funding |  | Limitations |
| --- | --- | --- |
| Most programs mention public funding combined with other types of fundraising. The funding occurred directly or by reimbursement and was strongly linked to universities. |  | Financial limitations caused the main impact on service since resources were smaller than demand. This factor limited the services and equipment maintenance offered. |
| Non-governmental organizations supported the programs, especially in countries with no government incentive. They were usually responsible for services not covered by health insurance. |  | Difficulties in professional recruitment and adherence were related to lower salaries for MDU dentists than for those in conventional practice and high demand for work. |
| Private initiatives were mainly constituted by sponsoring companies using social policies. |  | Patient recruitment and adherence were limitations generally related to the difficulty of disclosing the service to the community, different lifestyles, and difficult access to patients who did not want to attend the service. |
| Type of modality | Patient profile | Services offered |
| Van was the most cited due to lower cost than bus, satisfactory space, and practicality. | School-aged children comprised the largest public attended due to the school health-related programs. | Restoration with amalgam or composite resin and atraumatic restoration were the most offered. |
|  | Generally, they had reimbursement from government insurance. |  |
| Considering that trailers are independent of vehicles, their maintenance has advantages, and they offer greater space. | Usually, adults received support from projects dedicated to vulnerable populations, including homeless, patients with disabilities, and immigrants. | Extraction was the second most offered service. Some units expanded minor surgery to other services, such as biopsy and frenectomy. |
| The mobile structure was used in non-terrestrial transportation settings and areas with limited mobility. Generally, tents covered the service equipment. | Older adults received assistance in long-term care facilities, municipal centers, or at home. Usually, they were bedridden patients with compromised health. | Sealants were widely used due to the low complexity of execution, easy acceptance by patients, and preventive nature. |
|  |  | Preventive actions through oral health education were part of the policy of the program and used when clinical services could not be offered. |

Source: the author, 2024.

## DISCUSSION

This review demonstrated the versatility of MDU, such as reaching remote areas and attending socioeconomically vulnerable populations, being an excellent strategy for expanding access to health services. For example, literature described a detailed support plan for children of immigrant farmworkers in Southern California (18) and assistance for the Indigenous population (19), patients with disabilities (20–23), homeless (20–22, 24), pregnant women (25, 26), and LGBTQIA+ (27). Similarly, Pietrokovski *et al.* defined institutionalized older adults, patients with disabilities, and vulnerable populations as those attended by an Israeli network (21). Despite being distinct groups, they can be integrated into a similar approach design when arranged in the same geopolitical context.

Regarding patient profiles, children were the most attended in studies, mainly because of patient empowerment. School-based mobile dental programs are a viable solution to physical, financial, and structural barriers to dental care access, and proper oral healthcare is essential for children since it lays the foundation for oral health and disease prevention throughout life (28–30). Also, 47% of the offered services were oral hygiene education, and 39% were preventive sealants, highlighting the importance of teaching and reinforcing proper oral health habits in childhood to prevent biofilm-related diseases and complications.

Most services had assistive modality; 73% were restorations of decayed teeth, and 65% were extractions. These findings may be due to population, socioeconomic, political, and geographical factors, including logistical and transportation challenges due to geographical isolation, limiting access to dental care, low-income levels, and insufficient health coverage (31–33). Also, the limited infrastructure of some MDU may restrict specialized services, and the offered services were mainly curative care strongly integrated with preventive activities (34).

Each modality had unique characteristics shaped by a specific context. In the United States, the state of Minnesota has vast rural areas, and many counties do not have a dentist. Thus, school-based mobile dental programs were used regardless of Medicaid insurance to provide services for children and pregnant women due to this fact combined with professional shortage (25). In Peru, although some of the largest villages have government-run medical units, they lack dental services, and an international philanthropic institution operates using boats, transporting complete teams to remote areas every three months (35). During the study period, this service attended to 672 patients over 28 days, with ages ranging from 5 to 77 years. Also, Australia faces limited dental care in remote areas, increasing tooth decay in rural children (19). A non-profit organization provided dental care to 3,407 patients across 88 remote communities using air transport, delivering a total of 27,897 services (19). In this sense, a non-profit organization provided dental care using air transport. Therefore, a strong connection was observed among modalities of MDU, patient profile, and geographical context.

From a planning perspective, MDU require technical organization even before the arrival of professionals and coordination to structure a team addressing the population needs. Mulligan *et al.* reported that a previous visit was conducted approximately one year before to engage with the population and select strategies to identify children with the worst oral health (7). For organization during the intervention, a report from the British Journal of Dentistry described the coordination of a queuing system, screening area, and treatment area inside the unit, while volunteers simultaneously disclosed the MDU in the territory (36).

The integration of professionals, especially during the screening, is also essential to strengthen the interprofessional working process and approximate them to different health needs of patients. From a professional perspective, the recruitment and retention of these professionals in the service remain challenging. In South Africa, 60% of students reported working up to ten hours per day, while 87% stated that they treated more than thirty patients per day (37).

In this context, the decision to use MDU should be based on a field study due to the difficulties associated with implementation and maintenance costs. Although portable dental equipment has a relatively low cost (US$ 15,000 to US$ 20,000), vans can exceed US$ 300,000 (38). Also, transporting dental equipment requires the maintenance of vehicles, instruments, and materials due to constant vibration and movement during transport. Considering that the cooling of equipment depends on electrical service, the summer heat requires the removal of temperature-sensitive equipment (20). In addition, difficult terrain in which units usually operate (e.g., rural settlements) often leads to malfunctions of equipment, such as handpieces, water cooling, and aspirator (39). These factors can result in lost workdays due to technical or mechanical problems, and services become dependent on such repairs.

Funding directly influences the limitations of MDU and types of provided services. Preventive services are usually cheaper and easier than restorative services, and many locations increase the complexity of the program due to travel time and different site requirements (40). Also, the lack of specific funding for the program hinders professional recruitment and adherence, leading to reliance on volunteer work and highlighting the need to expand MDU services for children who are not eligible for government-funded dental insurance and other marginalized groups (7, 26).

Despite the challenges and recent implementation, MDU provide unique and innovative benefits regardless of territory. In India, 334 patients were treated in a three-week clinical period in 2009 (41). Also, the oral health-related quality of life improved by 65% for Brazilian children and 64.3% for their caregivers⁴¹. The study by Mickenautsch *et al.*, (39) performed in South Africa, reported that fewer teeth were extracted within a year of introducing atraumatic restorative treatment using MDU. Thus, the positive impacts of MDU justify its implementation, and adequate planning may improve the longevity of the program and its benefits.

The limitations of the review are related to the availability of studies that provide detailed information on this type of care. Many studies did not provide data on financing, service longevity or frequency of actions. Another point is the lack of use of validated instruments to assess the quality and effectiveness of the service offered.

This review included an innovative approach to understanding the organizational aspects of MDU services worldwide, focusing on structure, services, and results in different regions. However, detailed local studies are needed to better understand the developed scenario. The complications associated with the operation of MDUs are specific and closely tied to the environments in which they are deployed, making the standardized maintenance of these units unfeasible. To ensure longevity of the service, field surveys or interviews with professionals and patients are essential, as they enable the development of a situational diagnosis and facilitate target budgetary decision-making.

## CONCLUSION

Data analysis demonstrated that MDU play a crucial role in expanding access to dental care, especially in geographically remote areas and for vulnerable populations. The diversity of modalities and implementation flexibility allowed adaptation to local needs, providing curative and preventive care. Although most studies involved restorative procedures and reliance on public and volunteer funding, evidence pointed to a significant improvement in dental care for the populations.

## DECLARATIONS

### Registration information

This review had its protocol registered after data analysis.

### Availability of data and materials

The datasets generated and/or analyzed during the current study are not publicly available due to privacy policies, but are available from the corresponding author upon reasonable request.

### Authors’ contributions

EDM, AKLM and SQMS established the research protocol, the search strategy and applied it to the databases. EDM and AKLM screened the retrieved articles and extracted the results. AKLM created Figure 1, while EDM created Figures 2 and 3. AGRCO guided the steps between the authors. All authors read and approved the final manuscript.

## Acknowledgements

Not applicable

